# Redefining Delayed Graft Function: From Binary to Dynamic Using a Trajectory-Based Model to Evaluate Renal Recovery After Kidney Transplantation

**DOI:** 10.64898/2026.10.04.26362581

**Authors:** Malik Muhammad Makki, Komal Kumar, Harutyun Davtyan, Jiro Kusakabe, Eduardo Fernandes, Antonio Pinna, Khaled Refaai

## Abstract

**Background:** Delayed graft function (DGF) is among the most used metrics in transplant medicine, yet its prevailing definition-dialysis requirement within the first seven postoperative days lacks granularity and fails to capture the dynamic nature of graft recovery. It reflects a treatment decision rather than graft biology, varies by centre practice, and offers no temporal information about the pace or pattern of recovery. This study proposes a trajectory-based framework using serial serum creatinine measurements across three biologically defined time windows to capture the dynamic nature of early renal recovery.

**Methods:** We retrospectively analysed 205 adult kidney transplant recipients at Cleveland Clinic Weston Hospital in 2025. Donor groups included living donors, donation after brain death (DBD), donation after circulatory death with normothermic regional perfusion (DCD-NRP), and DCD without NRP. Creatinine slopes were estimated for Days 1-7, 7-30, and 30-90 using ordinary least-squares regression. Logistic regression assessed the predictive performance of each trajectory window for DGF, adjusting for Kidney Donor Profile Index, cold ischemia time, BMI, and donor type.

**Results:** DCD non-NRP kidneys showed negligible early creatinine decline (slope +0.02) compared to DBD (-0.71) and DCD-NRP (-0.57) grafts during Days 1-7. During Days 7-30, DCD-NRP kidneys showed accelerated recovery and converged toward DBD-level trajectories, whereas DCD non-NRP grafts remained sluggish. Predictive modelling identified the Day 7-30 trajectory had the strongest discrimination for DGF, with AUC 0.83, pseudo-R2 0.25, and the lowest AIC (181.73), outperforming Days 1-7 (AUC 0.75) and 30-90 (AUC 0.79). DGF rates were 0% in living donors, 25.5% in DBD, 20.7% in DCD-NRP, and 58.9% in DCD non-NRP recipients.

**Conclusions:** A trajectory-based creatinine model provides more informative prognostic value than the conventional binary DGF definition The Day 7-30 window periods offers the highest discriminatory power, and normothermic regional perfusion appears to favourably shift DCD kidney recovery trajectories in a manner the binary definition fails to detect.

## Introduction

Delayed graft function (DGF) is among the most widely reported endpoints in transplantation, yet its conceptual framework has remained largely static. As defined by the United Network for Organ Sharing (UNOS) and the Organ Procurement and Transplantation Network (OPTN), DGF is based on the requirement for dialysis within the first seven days post-transplant. Although simple and pragmatic, this binary definition lacks granularity and does not adequately reflect the continuum of early graft function and recovery. ^1^

This definition has shaped outcome reporting, program benchmarking, and clinical trial design across transplantation. While its simplicity is appealing, it comes at the expense of biological nuance. Reducing a complex and dynamic process to a binary outcome—dialysis within a fixed time window—fails to capture the heterogeneity of early graft recovery and injury. DGF has therefore been increasingly recognized as an imperfect surrogate for graft biology.^1–3^

Several limitations have been recognized. First, dialysis is a treatment decision, not an objective marker of graft biology; it reflects physician threshold, center culture, and fluid management protocols as much as it reflects underlying renal function.^2^ Second, the seven-day cutoff is retrospective by design—a graft cannot be classified until after the window has closed, limiting early intervention. Third, grafts with slow but uninterrupted recovery that never require dialysis may be just as impaired as those that do; the concept of “slow graft function” has been proposed to capture this intermediate phenotype. ^3^ Fourth, the emergence of DCD with normothermic regional perfusion (NRP) has created donor categories whose ischemia-reperfusion physiology may differ substantially from those encountered when the binary definition was established. ^4^

Creatinine kinetics may better capture the biological continuum of renal recovery. Serial creatinine trajectories have been proposed as a more dynamic framework for kidney graft assessment, and our group has recently reported that trajectory-based modelling may improve discrimination of delayed recovery patterns after kidney transplantation. ^11,13,17^ This approach is also conceptually aligned with recent mechanistic and clinical literature showing that ischemia-reperfusion injury, tubular repair, and early graft recovery unfold over time rather than as a single event.^4,14,16^

In this context, we propose a dynamic, trajectory-based approach to characterizing early graft recovery. Rather than asking whether dialysis occurred in week one, this model interrogates how creatinine moves across three biologically meaningful phases: Days 1–7, reflecting acute ischemia reversal; Days 7–30, corresponding to tubular repair and stabilization; and Days 30–90, indicating long-term trajectory. By fitting least-squares regression slopes to serial creatinine values within each window and incorporating these slopes into logistic regression models, we sought to determine which phase provides the strongest predictive signal for DGF, and how trajectory patterns differ across donor types.

## Methods

### Study Design and Population

This was a single-center, retrospective cohort study of adult kidney transplant recipients treated at Cleveland Clinic Weston Hospital in 2025. A total of 205 patients were included and stratified by donor type: living donors (n = 22), DBD (n = 98), DCD with NRP (n = 29), and DCD without NRP (n = 56). Standard clinical and laboratory data were extracted from the electronic medical record.

### Creatinine Trajectory Modelling

Serum creatinine values were collected at routine intervals during the first 90 postoperative days. For each patient and each phase, a slope was estimated using ordinary least-squares regression fitted to all available creatinine measurements within that window. Three trajectory phases were defined: Trajectory 1 (Days 1–7), hypothesized to reflect ischemic perfusion reversal; Trajectory 2 (Days 7–30), capturing intermediate tubular repair; and Trajectory 3 (Days 30–90), indicating long-term functional stabilization or progressive dysfunction. A negative slope denotes improving renal function; a positive slope denotes worsening or stagnation.

### Statistical Analysis

Logistic regression models were constructed separately for each trajectory window, with DGF (binary: dialysis within seven days) as the outcome. Each model was adjusted for KDPI, cold ischemia time, BMI, and donor type. Model performance was evaluated using the area under the receiver-operating characteristic curve (AUC), Pseudo R^2^ (Nagelkerke), and Akaike Information Criterion (AIC). Lower AIC values indicate better model fit; higher AUC values indicate stronger discrimination.

## Results

### Baseline Characteristics

Table 1 presents baseline characteristics stratified by donor type. Recipients of DCD non-NRP kidneys were oldest on average (mean 61.9 years) and had the highest KDPI (65.2 ± 20.6), consistent with marginal donor selection. Living donor recipients were youngest and had the lowest KDPI (50.0 ± 27.2). DGF occurred in none of the living donor recipients, 25.5% of DBD, 20.7% of DCD-NRP, and 58.9% of DCD non-NRP recipients.

**Table 1.**
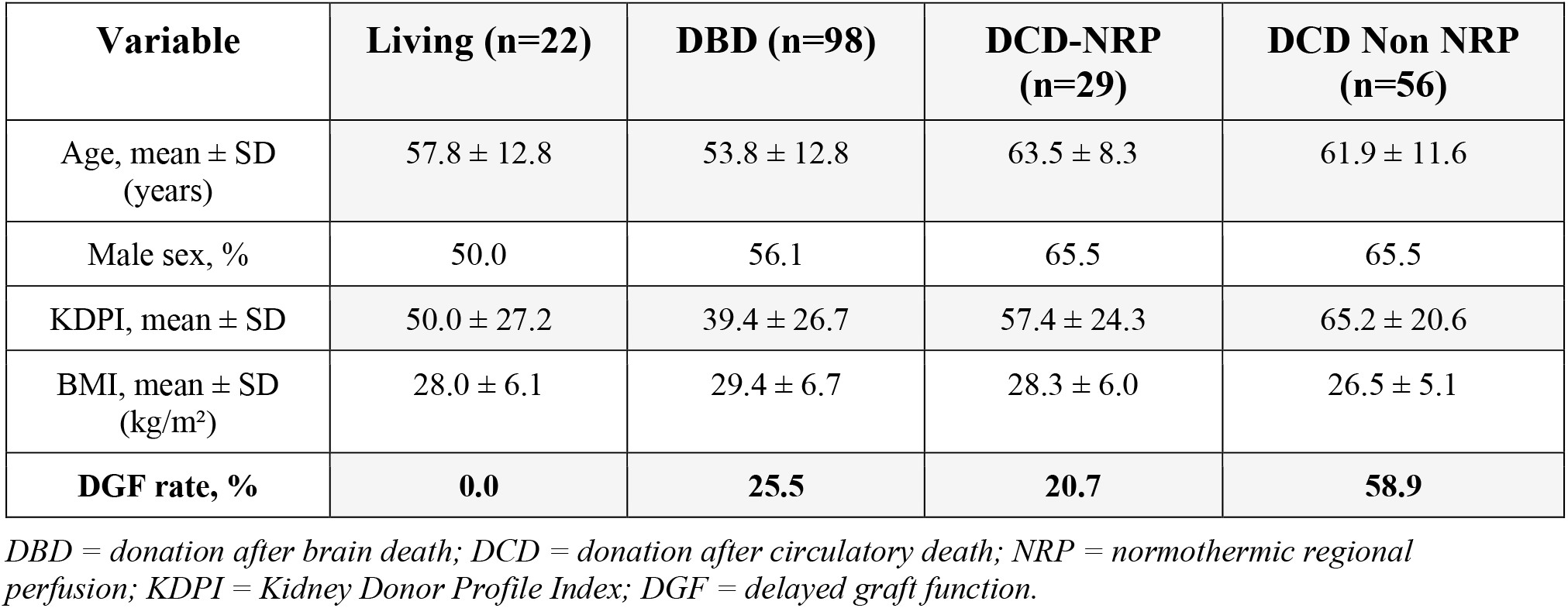
Baseline Characteristics of Kidney Transplant Recipients by Donor Type.

### Overall Creatinine Trajectory (Days 1–90)

Figure 1 displays the overall creatinine trajectory across all donor types from Day 1 through Day 90. Living donor recipients demonstrated the most rapid decline, reaching near-normal values by Day 3. DBD and DCD-NRP grafts followed intermediate trajectories with convergence by Day 30. DCD non-NRP grafts remained substantially elevated throughout the early period and exhibited the slowest and most heterogeneous descent.

**Figure 1.**
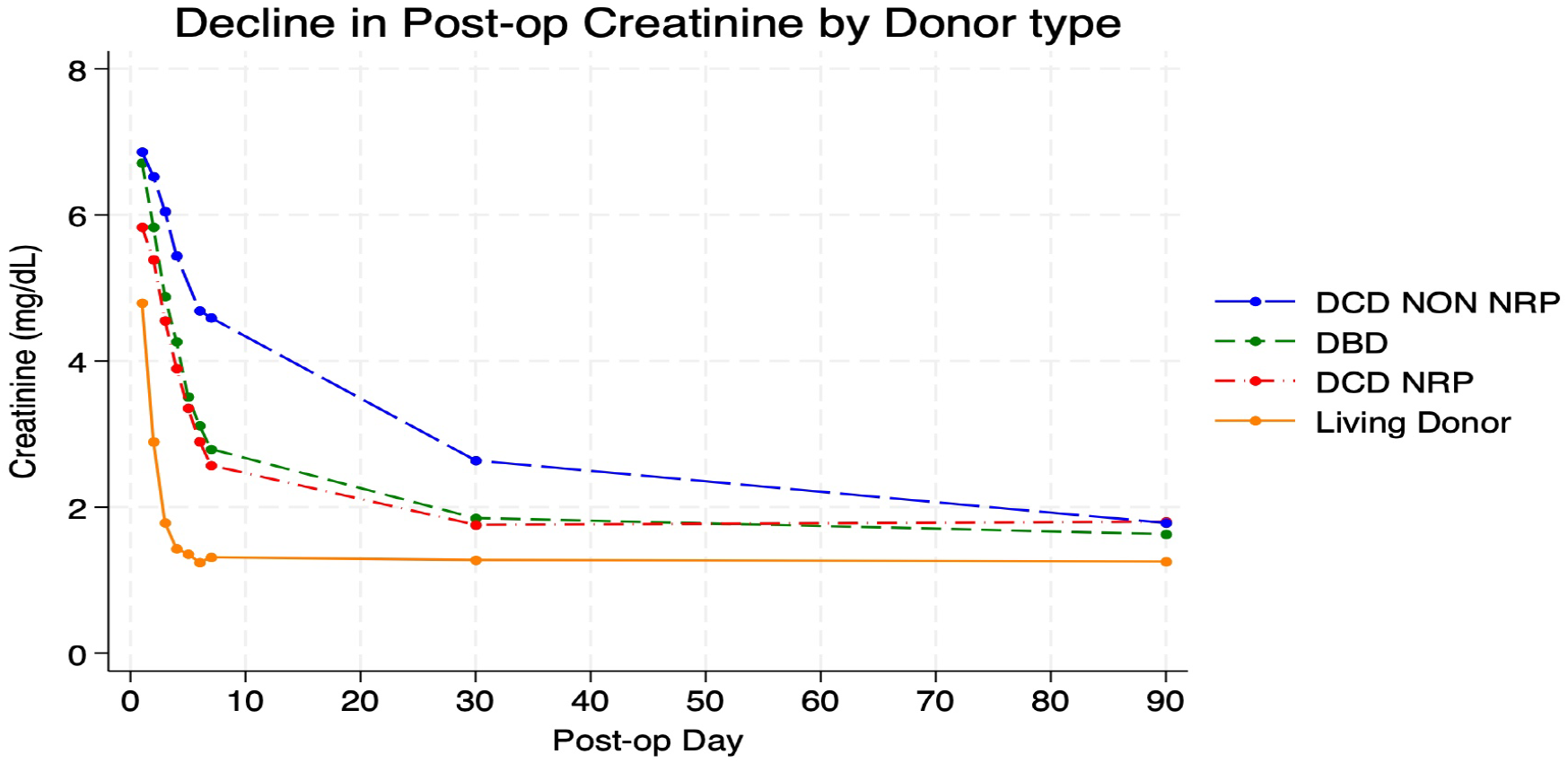
Overall Creatinine Trajectory by Donor Type (Days 1–90). Mean serum creatinine (mg/dL) plotted against post-operative day for all four donor categories. Living donors (orange) demonstrate the fastest recovery. DCD non-NRP kidneys (blue, dashed) remain elevated throughout the observation period. DBD (green) and DCD-NRP (red) trajectories converge by Day 30. DCD = donation after circulatory death; DBD = donation after brain death; NRP = normothermic regional perfusion.

### Trajectory 1: Days 1–7

During Days 1–7, DBD grafts exhibited the steepest creatinine decline (slope −0.71 mg/dL/day), consistent with rapid restoration of perfusion and early tubular recovery (Figure 2). DCD-NRP grafts showed a moderately steep decline (−0.57), while DCD non-NRP grafts demonstrated negligible change (+0.02), suggesting delayed or incomplete reversal of ischemia-reperfusion injury. Figure 5 further illustrates this divergence when stratified by DGF status: non-DGF grafts from all donor types show orderly parallel decline, while DGF grafts—particularly DCD non-NRP and DBD—maintain persistently elevated creatinine values through Day 7.

**Figure 2.**
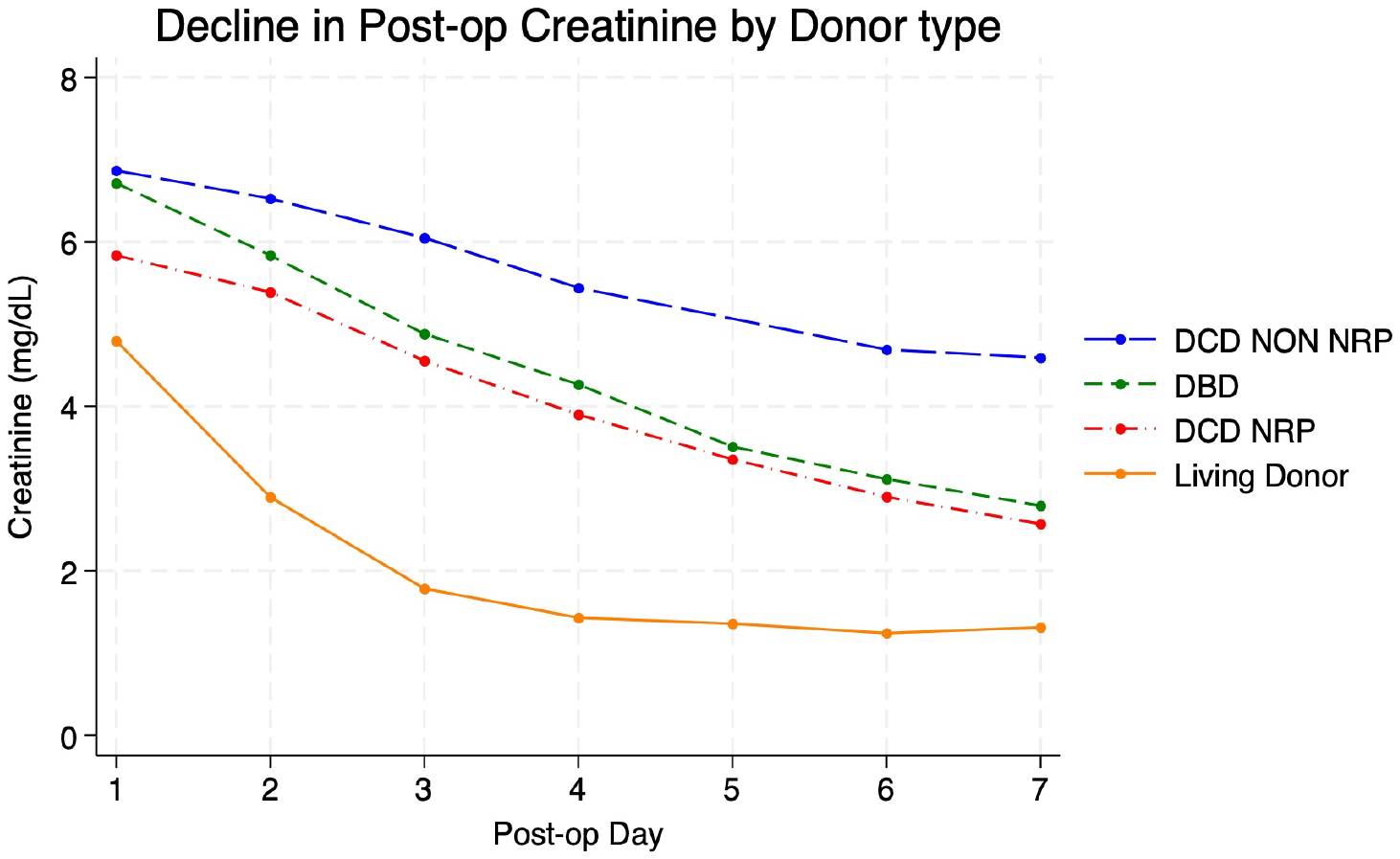
Creatinine Trajectory by Donor Type:Days 1–7. Mean serum creatinine (mg/dL) from post-operative Day 1 through Day 7. DBD grafts show the steepest decline (slope −0.71 mg/dL/day), followed by DCD-NRP (−0.57). DCD non-NRP grafts demonstrate negligible early recovery (slope +0.02). Living donors achieve near-normal values by Day 3.

### Trajectory 2: Days 7–30

Between Days 7 and 30, DCD-NRP grafts showed accelerated recovery (−0.035 mg/dL/day) and converged closely with the DBD trajectory (Figure 3). DCD non-NRP grafts recovered more slowly and with greater variability (−0.087). When stratified by DGF status (Figure 6), DGF recipients from all three deceased-donor groups displayed ongoing steep creatinine decline during this window, highlighting the continued biological relevance of the intermediate phase even in grafts that required dialysis.

**Figure 3.**
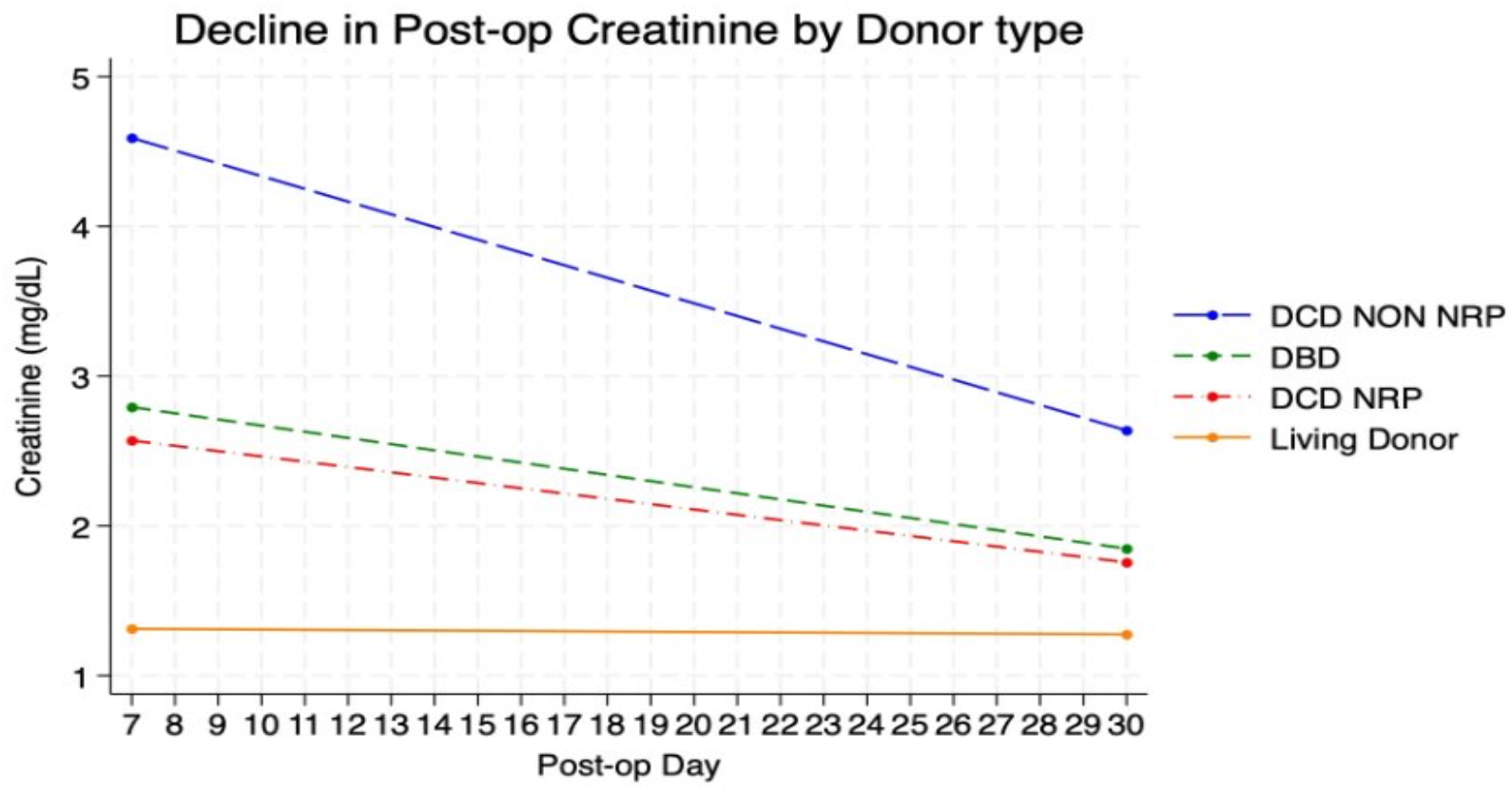
Creatinine Trajectory by Donor Type: Days 7–30. Mean serum creatinine (mg/dL) from post-operative Day 7 through Day 30. DCD-NRP grafts accelerate recovery during this phase, converging with DBD trajectories. DCD non-NRP grafts recover more slowly and with greater variability. Living donors maintain near-normal and stable creatinine.

### Trajectory 3: Days 30–90

During Days 30–90, DBD and DCD-NRP grafts stabilized near-normal creatinine values, while DCD non-NRP grafts remained heterogeneous (Figure 4). Figure 7 shows that among non-DGF recipients, trajectories are largely flat and stable. Among DGF recipients, DCD-NRP grafts maintained the lowest creatinine values, while DCD non-NRP and DBD DGF grafts continued a gradual decline from elevated baselines.

**Figure 4.**
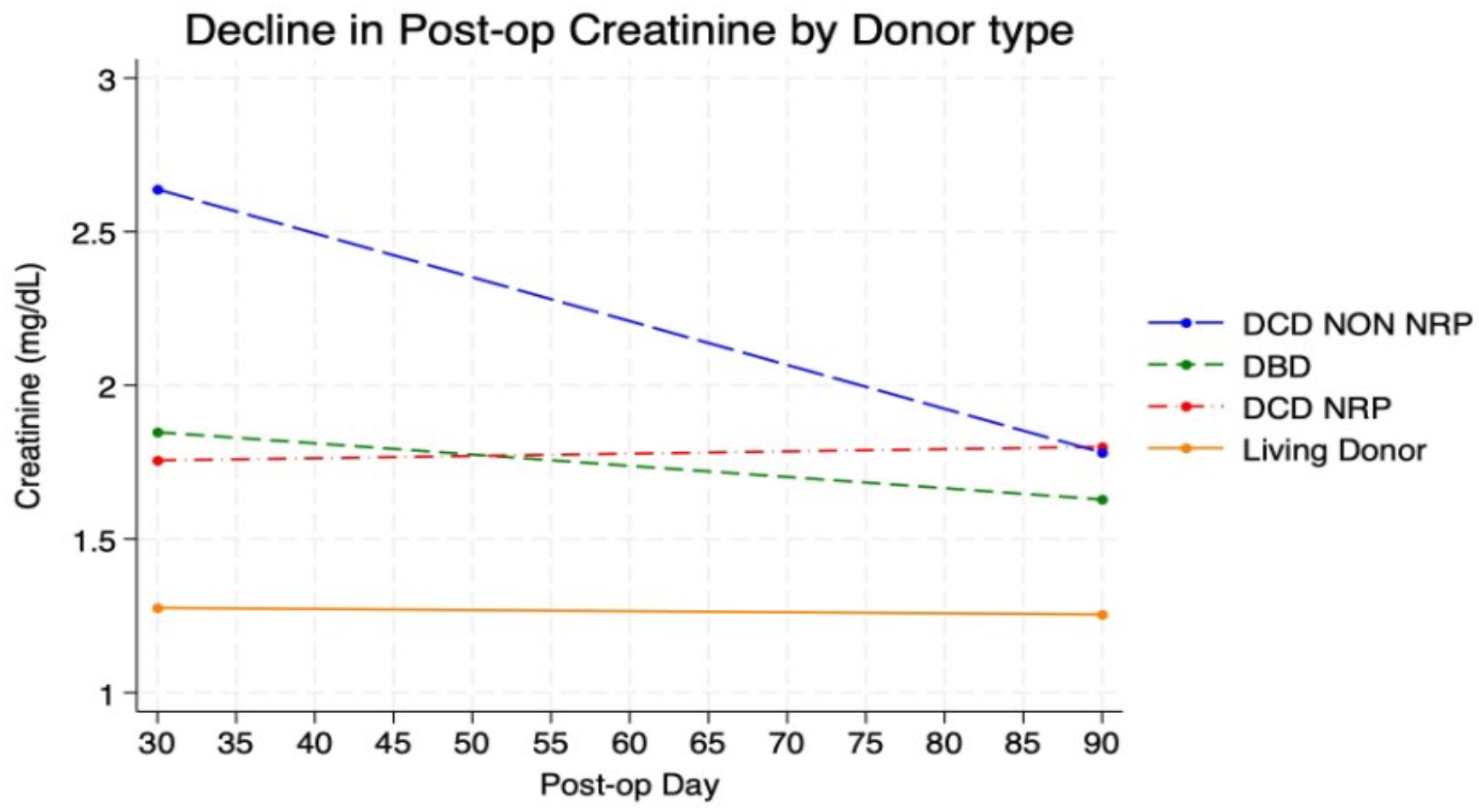
Creatinine Trajectory by Donor Type: Days 30–90. Mean serum creatinine (mg/dL) from post-operative Day 30 through Day 90. DBD and DCD-NRP grafts stabilize near-normal values. DCD non-NRP grafts remain elevated and heterogeneous, with a subset demonstrating persistent creatinine elevation suggestive of early chronic allograft injury.

**Figure 5.**
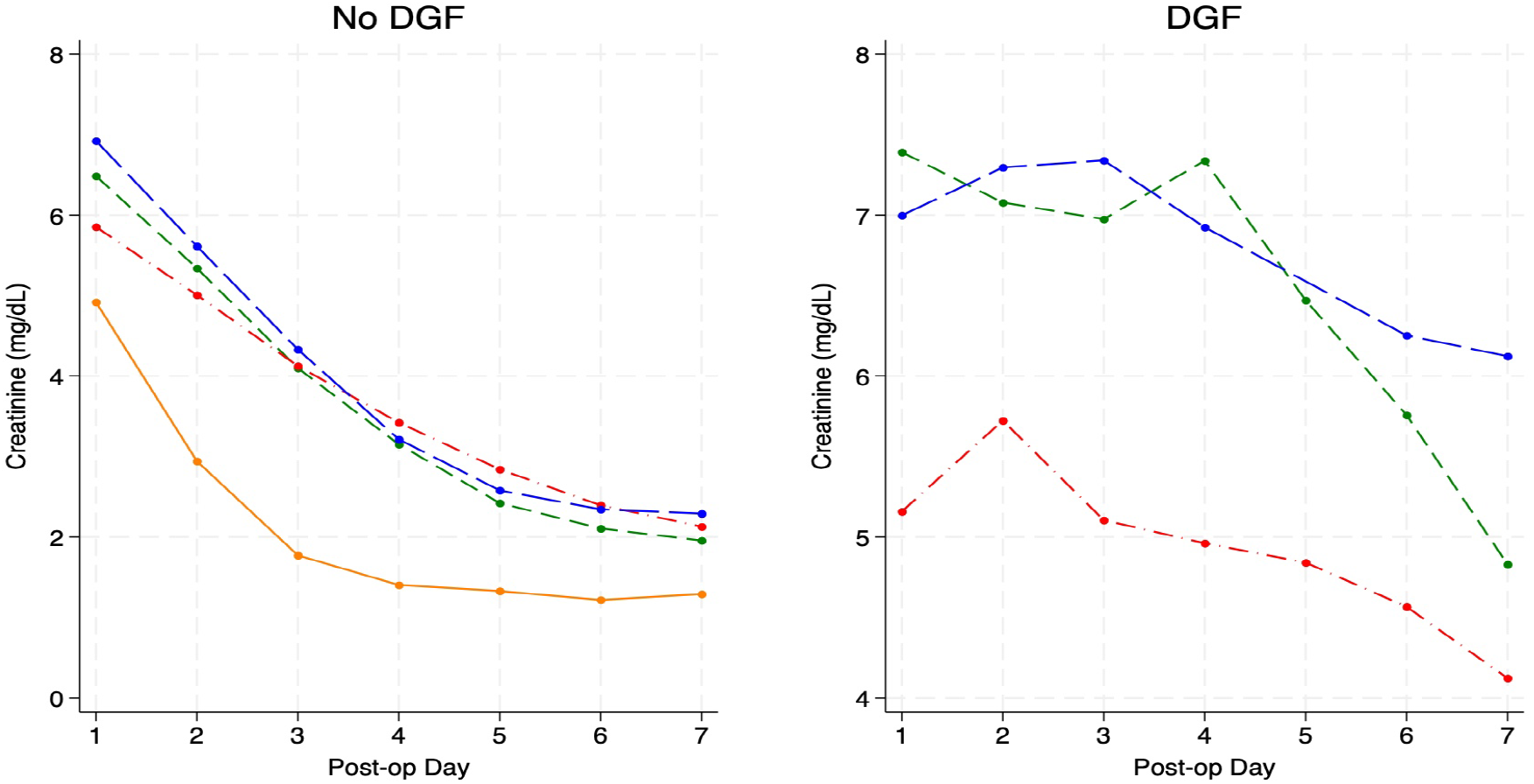
Creatinine Trajectory Days 1–7 Stratified by DGF Status. Left panel shows trajectories in non-DGF recipients across donor types; right panel shows DGF recipients. Non-DGF grafts demonstrate orderly parallel decline. DGF grafts, particularly DCD non-NRP (blue) and DBD (green), sustain markedly elevated creatinine throughout Days 1–7, reflecting ongoing ischemic injury.

**Figure 6.**
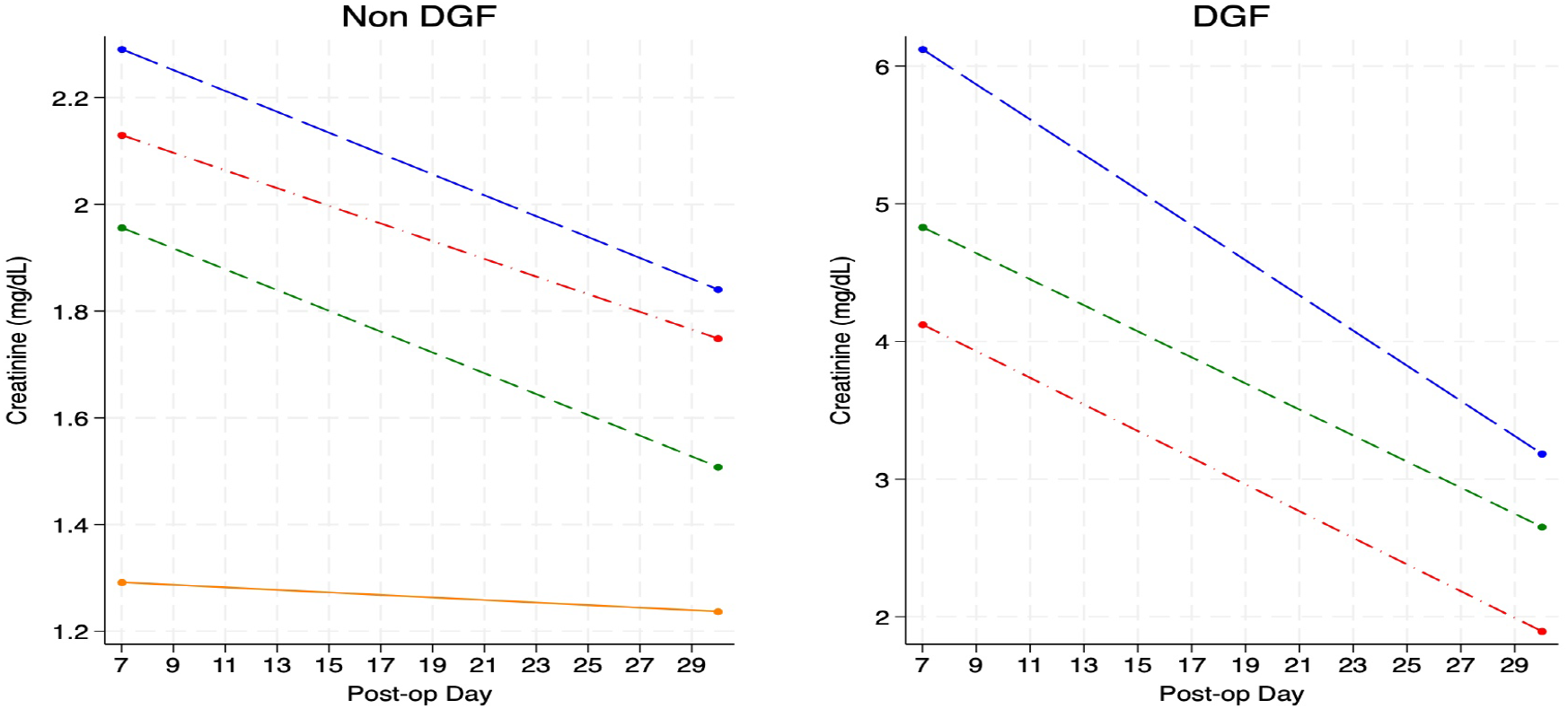
Creatinine Trajectory Days 7–30 Stratified by DGF Status. Left panel: non-DGF recipients show flat to mildly declining trajectories consistent with stabilized function. Right panel: DGF recipients demonstrate continued steep creatinine descent, reflecting ongoing but belated tubular recovery. DCD-NRP (red) DGF grafts show the most rapid recovery among the DGF cohort during this window.

**Figure 7.**
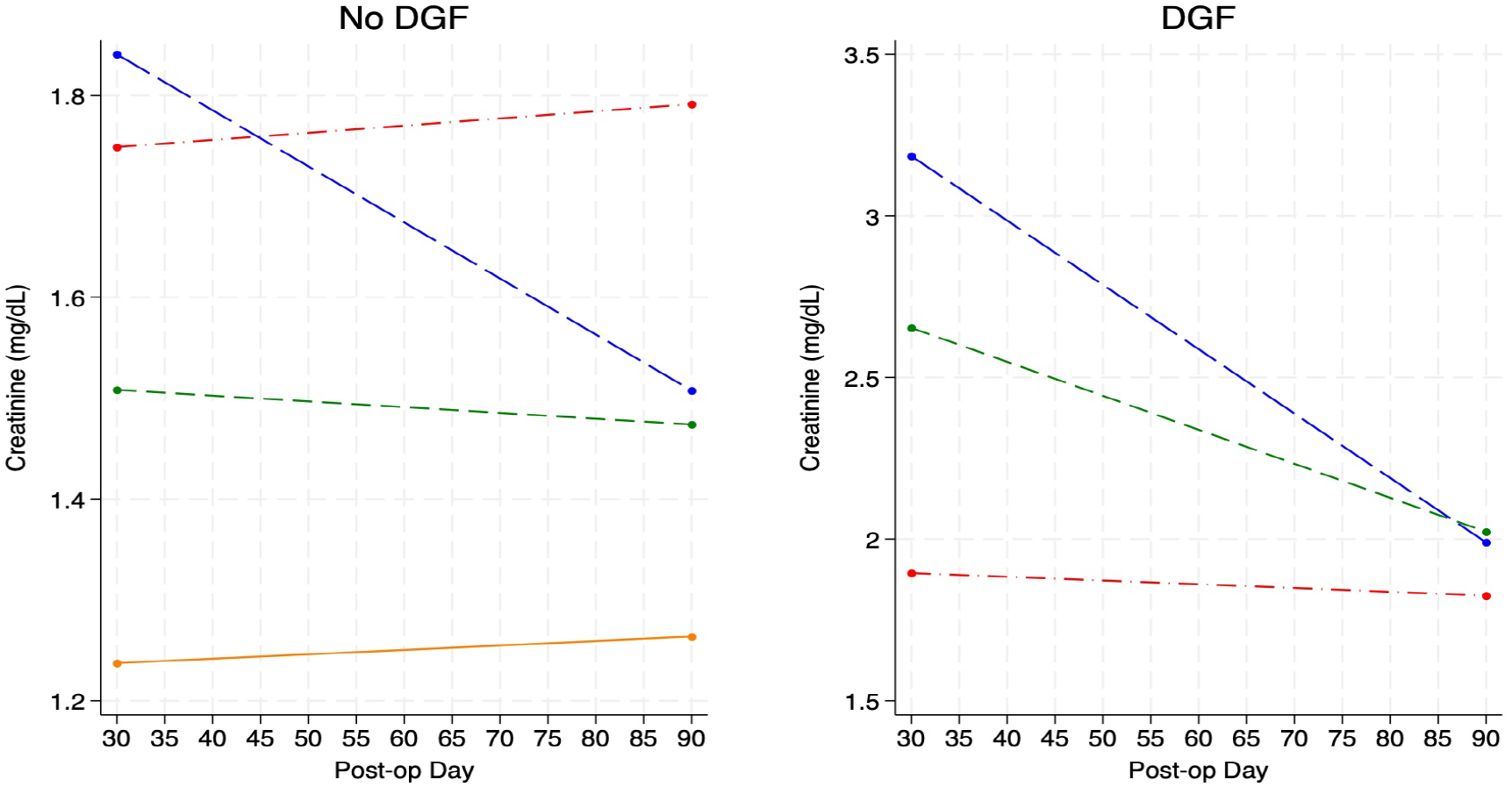
Creatinine Trajectory Days 30–90 Stratified by DGF Status. Left panel (non-DGF): all donor types show stable or near-flat trajectories. Right panel (DGF): DCD-NRP grafts (red) maintain the lowest values. DCD non-NRP (blue) and DBD (green) DGF grafts continue gradual decline from elevated starting points, reflecting delayed functional consolidation.

### Predictive Model Performance

Table 2 summarizes the discriminatory performance of creatinine slope models across the three trajectory windows. The Day 7–30 slope yielded the strongest discrimination (AUC 0.83), the highest explanatory power (Pseudo R^2^ 0.25), and the best model fit by AIC (181.73). The Day 1– 7 slope performed modestly well (AUC 0.75), while the Day 30–90 slope showed intermediate performance (AUC 0.79).

**Table 2.**
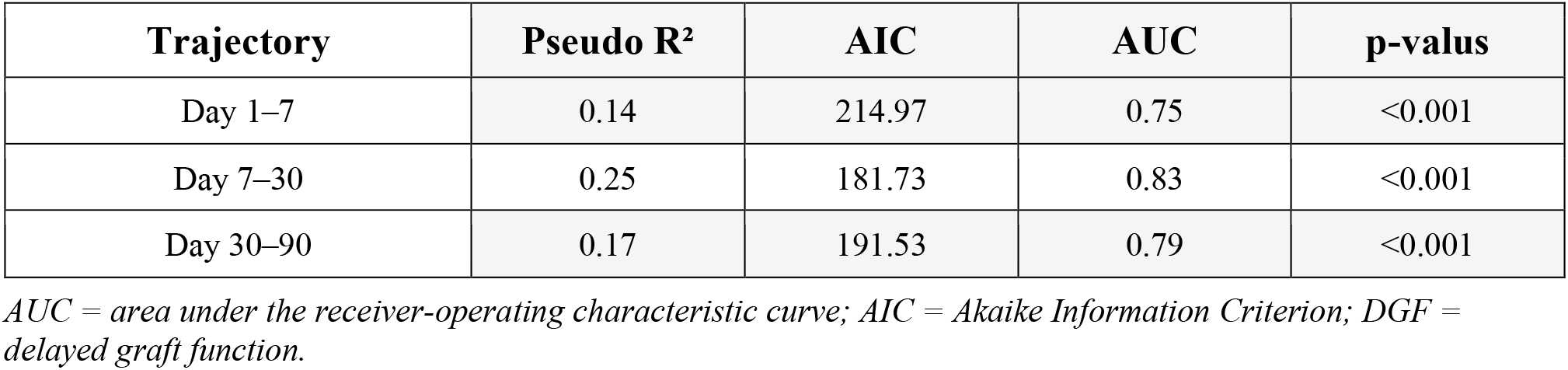
Predictive Performance of Creatinine Trajectory-Based Models for DGF.

| Trajectory | Pseudo $R^2$ | AIC | AUC | p-value |
| --- | --- | --- | --- | --- |
| Day 1–7 | 0.14 | 214.97 | 0.75 | <0.001 |
| Day 7–30 | 0.25 | 181.73 | 0.83 | <0.001 |
| Day 30–90 | 0.17 | 191.53 | 0.79 | <0.001 |

AUC = area under the receiver-operating characteristic curve; AIC = Akaike Information Criterion; DGF = delayed graft function.

Predicted probability curves for DGF illustrated donor-specific risk profiles across each phase (Figures 8–9). In the Day 1–7 window (Figure 8), DCD non-NRP kidneys showed the steepest increase in DGF probability as creatinine slope worsened, indicating heightened sensitivity to early ischemic injury. DCD-NRP grafts showed a right-shifted curve, suggesting NRP confers immediate protective effects that lower DGF probability even when early creatinine slopes are poor.

**Figure 8.**
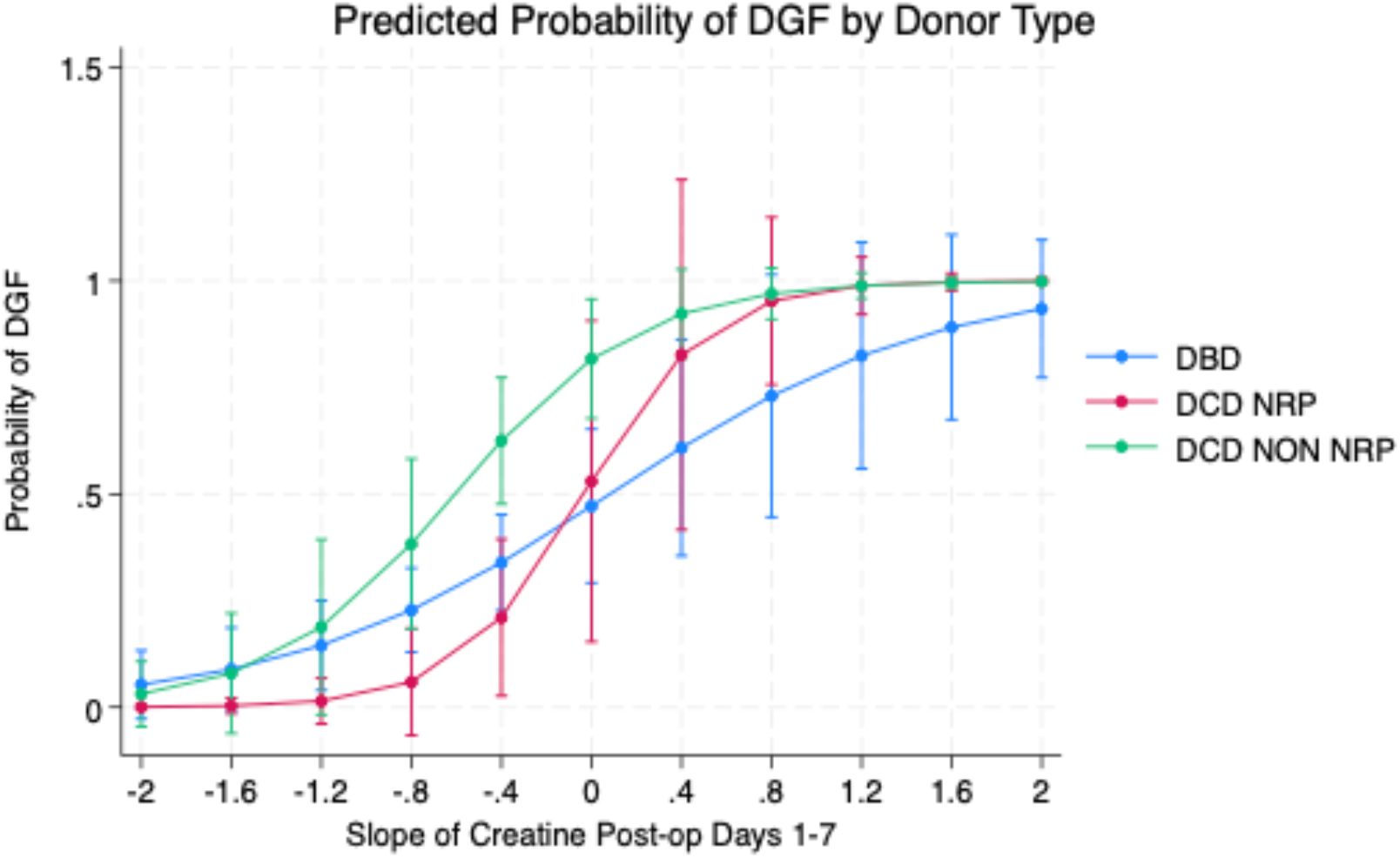
Predicted Probability of DGF by Creatinine Slope: Days 1–7. Logistic regression–derived probability of DGF as a function of the Day 1–7 creatinine slope by donor type. DCD non-NRP (green) shows the steepest risk increase with worsening slope. DCD-NRP (red) curve is right-shifted, indicating NRP attenuates early DGF risk. DBD (blue) demonstrates intermediate susceptibility.

In the Day 7–30 window (Figure 9), DCD-NRP grafts demonstrated the fastest decline in DGF probability as creatinine trajectories improved, consistent with more effective late-phase tubular repair. In the Day 30–90 window (Figure 10), the DGF signal in DCD-NRP kidneys was consistently low and flat, reflecting durable recovery irrespective of late creatinine dynamics.

**Figure 9.**
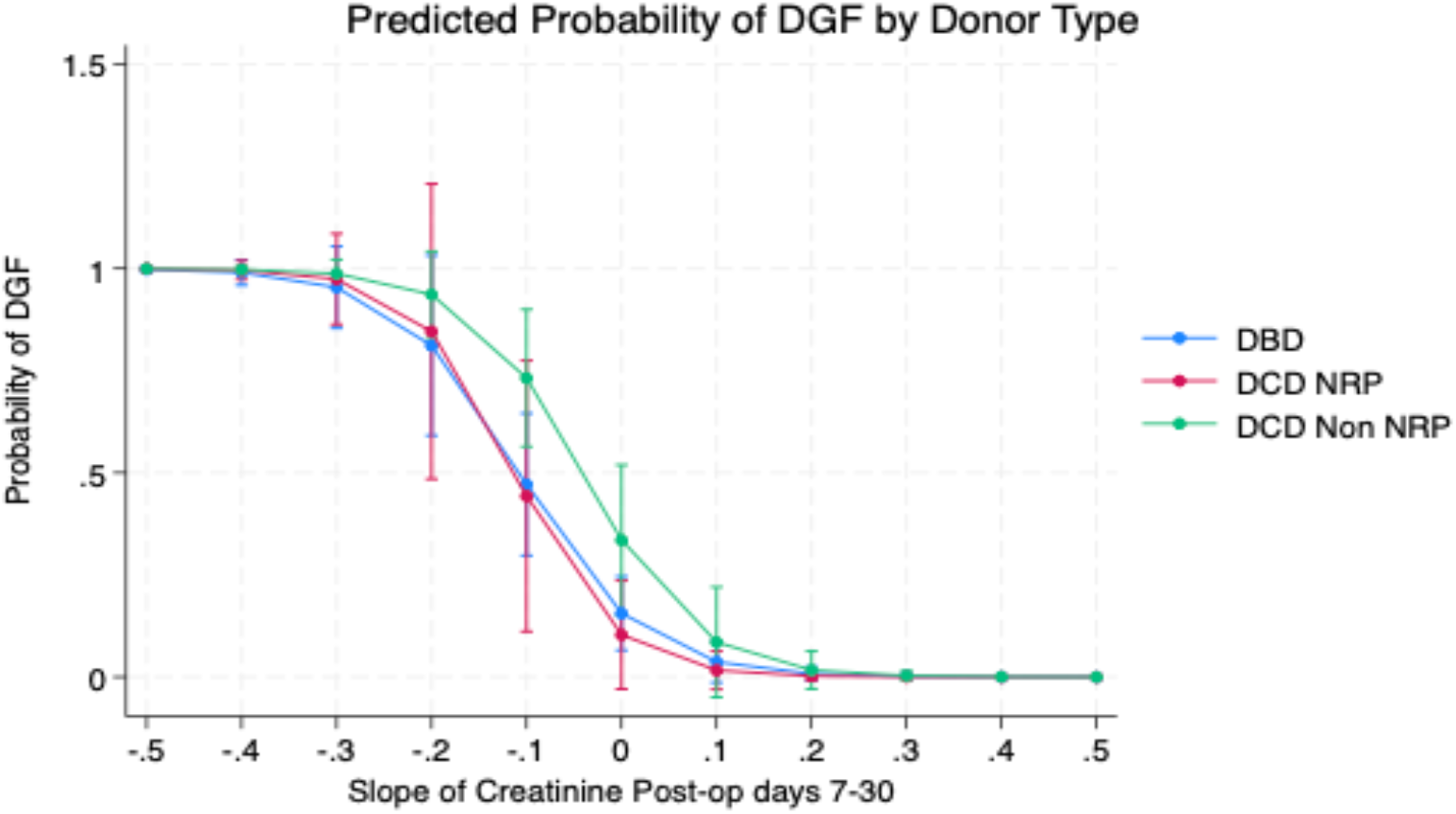
Predicted Probability of DGF by Creatinine Slope: Days 7–30. Logistic regression–derived probability of DGF as a function of the Day 7–30 creatinine slope by donor type. The Day 7–30 window provides the strongest model discrimination overall (AUC 0.83). DCD-NRP (red) demonstrates the fastest risk reduction as recovery slope improves, reflecting more effective intermediate-phase tubular repair.

**Figure 10.**
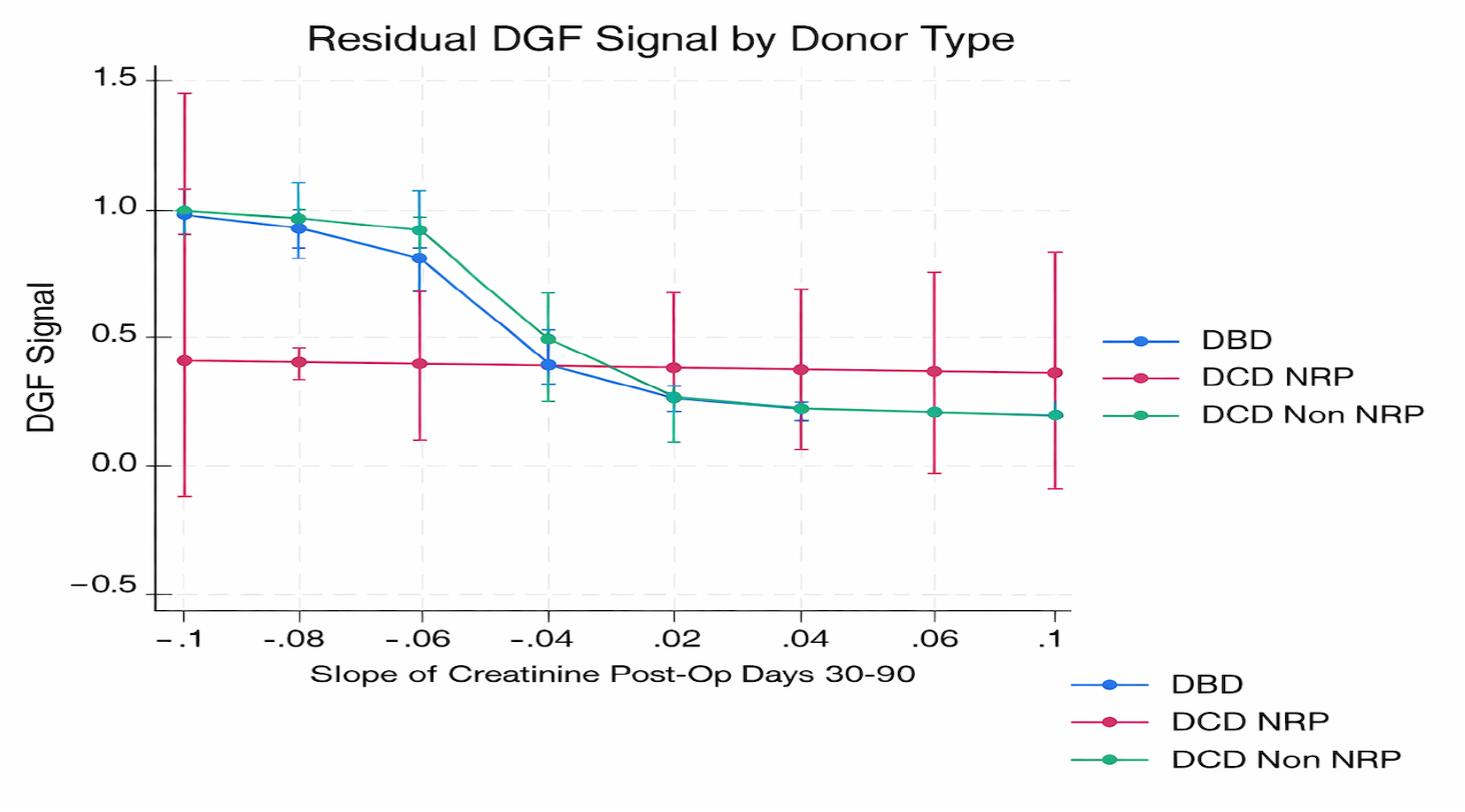
Residual DGF Signal by Creatinine Slope: Days 30–90. Post-hoc DGF signal plotted against the Day 30–90 creatinine slope. DCD-NRP (red) demonstrates a consistently low, flat signal irrespective of late creatinine dynamics, confirming durable recovery. DBD (blue) signal declines rapidly with improvement. DCD non-NRP (green) shows gradual, heterogeneous signal reduction.

## Discussion

This study makes a conceptually straightforward but practically significant argument: graft recovery is a process, not a moment, and measuring it as one requires a framework that tracks how function evolves over time. The trajectory-based model we propose does not require new data collection; it reframes existing creatinine values—already measured in every transplant recipient—as a dynamic biological signal.

The days 7–30 period emerged as the most clinically meaningful window to assess graft recovery. Early post-transplant function is often dominated by transient factors—ischemia–reperfusion injury, surgical variables, and early management decisions—that obscure the true biological signal. By the second week, as these influences diminish, the trajectory of kidney function begins to reflect the graft’s inherent recovery potential more reliably. This is consistent with prior work showing that early creatinine kinetics and slow graft function are prognostically important.^2,3,11–13^

The differential performance of DCD-NRP versus DCD non-NRP kidneys across all three phases is particularly noteworthy. Although both donor categories share similar ischemic mechanisms at the point of cardiac arrest but diverge substantially in their functional trajectories after transplantation. NRP-treated grafts converging toward DBD-level recovery by Day 30. This finding corroborates emerging evidence suggesting that NRP mitigates ischemia-reperfusion injury through mechanisms including active cellular repair during the perfusion period and improved mitochondrial function in the recovered organ.^5-8^

In a large propensity-matched multicentre study, Padilla et al. demonstrated that rapid recovery without NRP was associated with nearly twice the odds of DGF compared to NRP in controlled DCD transplants. ^6^ Oniscu et al., using national registry data from the United Kingdom, similarly found that NRP kidneys had 35% lower adjusted odds of delayed graft.

At present, DGF-attributed outcomes—including higher acute rejection rates, increased graft failure, lower one-year eGFR, and prolonged hospitalization—are derived largely from studies using the binary dialysis-based definition.^8^ A 2023 meta-analysis confirmed that DGF is associated with significantly worse short- and long-term outcomes, including a nearly twofold increase in acute rejection risk and meaningfully lower one-year eGFR.^9^ If trajectory analysis reclassifies a proportion of dialyzed grafts as intermediate recoverers rather than truly delayed ones, the magnitude of adverse outcomes traditionally attributed to DGF may shift. Conversely, slowly recovering non-dialyzed grafts that are currently invisible to the binary metric may harbor a disproportionate share of long-term risk; emerging data suggest slow graft function independently predicts graft survival loss across multiple donor types.^10^

This study has several limitations. The retrospective, single-center design limits generalizability and introduces the risk of unmeasured confounding. The sample size, particularly in the DCD-NRP subgroup, constrains the stability of subgroup estimates. Creatinine trajectories may be influenced by clinical decisions such as diuretic use and fluid management protocols. Patients who required dialysis for DGF represent a specific challenge: dialysis itself alters creatinine dynamics and may introduce systematic bias in slope estimation. Future work should address these limitations through prospective, multicenter cohorts with standardized creatinine collection protocols and functional biomarker integration.

Nevertheless, the findings support a more nuanced conceptualization of early kidney graft recovery. The Days 7–30 interval appears to be the most informative phase for predicting DGF, and NRP appears to shift DCD recovery trajectories in a favourable direction. These observations suggest that dynamic biomarkers may improve risk stratification and may eventually inform more individualized post-transplant management.^4–8,11–16,17^

## Conclusions

Delayed graft function, as currently defined, simplifies a complex and heterogeneous biological process into a binary label. A trajectory-based approach using serial creatinine measurements across post-transplant phases provides a clinically meaningful assessment of graft recovery. The Days 7–30 window demonstrate the greatest discriminatory value for DGF, reflecting intrinsic graft function after early perioperative influences subside. Importantly, normothermic regional perfusion (NRP) favourably modifies DCD kidney recovery trajectories—an effect not captured by conventional definitions.

Adopting dynamic recovery metrics may improve risk stratification, program benchmarking, and guide more tailored post-transplant management strategies

## Data Availability

The datasets generated and analysed during this study are not publicly available due to patient privacy restrictions but are available from the corresponding author on reasonable request, subject to institutional approval.

## Ethics Statement

This retrospective single-center study was reviewed and approved by the Cleveland Clinic Weston Hospital Institutional Review Board. The requirement for informed consent was waived because the study used previously collected clinical data and posed minimal risk to participants.

## References

1. Mallon DH, Bradley JA, Watson CJE, Pettigrew GJ. Delayed graft function: state of the art, expert perspective, and anticipated progress over the next decade. Am J Transplant. 2013;13(5):1130–1141. doi:10.1111/ajt.12142.

2. Fehrenbach DA, Bonventre JV. Mechanisms of maladaptive repair after acute kidney injury leading to accelerated kidney ageing and chronic kidney disease. Nat Rev Nephrol. 2015;11(5):264–276.

3. Cohen JB, Reese PP. Association of slow graft function with long-term outcomes in kidney transplant recipients. Am J Transplant. 2018;18(12):2948–2957. doi:10.1111/ajt.14985.

4. Salvado E, Damiano I, Ballarini G, et al. Ischemia-reperfusion injury in kidney transplantation: mechanisms and potential therapeutic targets. J Clin Med. 2024;13(8):2284. doi:10.3390/jcm13082284.

5. Padilla M, Coll E, Fernández-Pérez C, et al. Improved short-term outcomes of kidney transplants in controlled donation after the circulatory determination of death with the use of normothermic regional perfusion. Am J Transplant. 2021;21(11):3618–3628. doi:10.1111/ajt.16579.

6. Oniscu GC, Mehew J, Butler AJ, et al. Improved organ utilization and better transplant outcomes within situ normothermic regional perfusion in controlled donation after circulatory death. Transplantation. 2023;107(2):438–448. doi:10.1097/TP.0000000000004280.

7. Klein Nulend R, Hameed A, Singla A, et al. Normothermic machine perfusion and normothermic regional perfusion of DCD kidneys before transplantation: a systematic review. Transplantation. 2024;108(11):2198–2209. doi:10.1097/TP.0000000000005150.

8. Hosgood SA, Callaghan CJ, Wilson CH, et al. Normothermic machine perfusion versus static cold storage in donation after circulatory death kidney transplantation: a randomized controlled trial. Nat Med. 2023; 29:1511–1519. doi:10.1038/s41591-023-02376-7.

9. Motter JD, Tangri N, Damrauer SM, et al. Effects of delayed graft function on transplant outcomes: a meta-analysis. Transplantation Direct. 2023;9(2):e1450. doi:10.1097/TXD.0000000000001450.

10. Husain SA, Roke MC, Mohan S, et al. Kidney transplant outcomes after prolonged delayed graft function. Transplantation. 2022;106(3):600–609. doi:10.1097/TP.0000000000003826.

11. Makki MM, Kumar K, Davtyan H, Fernandes E, Kusakabe J, Pinna A, Refaai K. Redefining delayed graft function: from binary to dynamic using a trajectory-based model to evaluate renal recovery after kidney transplantation. Am J Transplant. 2026;26(1 Suppl 1): S11–S12. doi: 10.1016/j.ajt.2025.12.038.

12. Makki MM, Kumar K, Davtyan H, Hussein A, Fernandes E, Refaai K. Donor KDPI and predicted risk of delayed graft function in NRP kidney transplant recipients. Am J Transplant. 2026;26(1 Suppl 1): S28. doi: 10.1016/j.ajt.2025.12.074.

13. Makki MM, Kumar K, Davtyan H, Hussein A, Kusakabe J, Fernandes E, Refaai K. Recipient body mass index and delayed graft function after normothermic regional perfusion. Am J Transplant. 2026;26(1 Suppl 1): S88. doi: 10.1016/j.ajt.2025.12.192.

14. Bhattacharjee RR, Major RWH, Moledina N. Early creatinine kinetics after kidney transplantation and its association with outcomes: a systematic review. Transplantation Reviews. 2022;36(2):100688. doi: 10.1016/j.trre.2022.100688.

15. Valentin MO, Holdaas H, Kubler B, et al. Clinical events and renal function in the first year predict long-term kidney transplant survival. Clin J Am Soc Nephrol. 2022;17(4):573–583. doi:10.2215/CJN.13671021.

16. Chen Y, Zou M, Zhang Y, et al. Risk factors for delayed graft function in patients with kidney transplantation: a systematic review and meta-analysis. Ren Fail. 2025;47(1):2467894. doi:10.1080/0886022X.2025.2467894.

17. Creatinine Trajectories: A Dynamic Predictor of Post-Kidney Transplant Recovery and Delayed Graft Function. Makki MM, Kumar K, Davtyan H, Kusakabe J, Fernandes E, Pinna A, Refaai K. J Urol. 2026;215(-[5S):e240. doi:10.1097/01.JU.0001191324.44573.90.29

